# The ChAMP App: A Scalable mHealth Technology for Detecting Digital Phenotypes of Early Childhood Mental Health

**DOI:** 10.1101/2023.01.19.23284753

**Authors:** Bryn C. Loftness, Julia Halvorson-Phelan, Aisling O’Leary, Carter Bradshaw, Shania Prytherch, Isabel Berman, John Torous, William L. Copeland, Nick Cheney, Ryan S. McGinnis, Ellen W. McGinnis

## Abstract

Childhood mental health problems are common, impairing, and can become chronic if left untreated. Children are not reliable reporters of their emotional and behavioral health, and caregivers often unintentionally under-or over-report child symptoms, making assessment challenging. Objective physiological and behavioral measures of emotional and behavioral health are emerging. However, these methods typically require specialized equipment and expertise in data and sensor engineering to administer and analyze. To address this challenge, we have developed the ChAMP (Childhood Assessment and Management of digital Phenotypes) System, which includes a mobile application for collecting movement and audio data during a battery of mood induction tasks and an open-source platform for extracting digital biomarkers. As proof of principle, we present ChAMP System data from 101 children 4-8 years old, with and without diagnosed mental health disorders. Machine learning models trained on these data detect the presence of specific disorders with 70-73% balanced accuracy, with similar results to clinical thresholds on established parent-report measures (63-82% balanced accuracy). Features favored in model architectures are described using Shapley Additive Explanations (SHAP). Canonical Correlation Analysis reveals moderate to strong associations between predictors of each disorder and associated symptom severity (r = .51-.83). The open-source ChAMP System provides clinically-relevant digital biomarkers that may later complement parent-report measures of emotional and behavioral health for detecting kids with underlying mental health conditions and lowers the barrier to entry for researchers interested in exploring digital phenotyping of childhood mental health.

## I. Introduction

CHILDHOOD mental health problems are common and can become chronic and impairing if left untreated. Children eight and younger are not reliable reporters of their emotional symptoms, making it particularly challenging to assess their emotional and behavioral health. Moreover, caregivers often unintentionally under-report their children’s emotional health, or can exhibit reporting bias based on their own mental health [1]. Because of these challenges in assessment, early childhood researchers have started to explore objective measures of emotional and behavioral health captured directly from a child’s physiology and behaviors.

Successful systems [2], [3] for capturing child physiology and behavior typically leverage mood induction tasks, or behavioral paradigms that “press” for a specific emotional or behavioral response [4]. A child’s response is quantified via a physiological metric (i.e., galvanic skin response, heart rate) measured using specialized equipment (e.g., [5], [6]) and/or via behavioral coding (i.e., tracking facial expressions/body movements in task videos [3], [7]) which requires specialized computer programs (e.g., [8]) and trained personnel [4], [9]. In some cases, quantifiable features responsive to the behavioral paradigms and related to the child’s emotional and behavioral health [10] are then used to supplement clinical assessment [4],[11], [12].

Despite the plethora of research (i.e., [4], [9], [10], [12]–[14]) and some clinical utility [4] of objective physiological and behavioral measures during mood induction tasks and their links with early childhood emotional and behavioral health, there are important challenges that limit wide use outside of the original authors’ laboratories. Most of these systems require a) specialized measurement equipment which may be expensive, require expert knowledge to use and interpret, and are cumbersome to wear for young children and/or b) video coding which require personnel effort, and time to train and code reliably. There is an unmet need for a system that can capture objective measures of a child’s physiological and behavioral response to mood inductions that is accessible for patients, researchers, and providers, and could enable the broad use, validation, and iterative development of these approaches.

Digital phenotyping, which leverages metrics available from wearable sensors and smartphones for detecting symptoms and presence of underlying health conditions, holds great promise for addressing this unmet need [15]. While there are some great early examples (e.g., smartphone metrics to detect autism in young children [16]), there is still much to learn about digital phenotypes, particularly in early childhood.

Our prior work has demonstrated that digital phenotyping derived from wearable sensor and audio data during mood induction paradigms can identify young children with internalizing disorders with good accuracy (75%-81%) [17]– [23]. However, it can be difficult for researchers to replicate this paradigm as an interdisciplinary team with expertise in wearable sensors, biomechanics, signal processing, data science, and clinical psychology is required to meet the important requirements for stringent validation [24]. We also note that these previous models did not classify specific diagnoses or severity, which are important outputs to consider in a future complementary clinical tool that may leverage similar data streams.

In this work, our team presents the ChAMP (Childhood Assessment and Management of digital Phenotypes) System, which is an accessible smartphone app and open-source platform for collecting and interpreting motion and audio data during mood induction tasks. In so doing, we aim to make standardized, validated digital phenotyping tools accessible to other researchers to enable efficacy and effectiveness testing across diverse populations and use-cases [15], [25], [26]. This approach ultimately allows evidence-based complementary digital mental health tools, such as this system and accompanying models, to get into the hands of patients and providers more swiftly. The technological advances and findings in this work build considerably on past work. Namely, we 1) collect a new sample of over 100 children and their families to evaluate our newly developed ChAMP System, 2) and explore its ability to detect specific diagnoses such as anxiety, depression, and ADHD via machine learning approaches deployed on the data it collects and 3) extend our past work evaluating individual mood-induction tasks by considering data from a combination of tasks and for predicting individual diagnoses, as opposed to general internalizing disorders.

## II. Methods

### A. The ChAMP System

The ChAMP System standardizes the administration of behavioral mood induction tasks for children (Table 1) by providing a repeatable and replicable digital framework, enabling collection of multimodal physiological data at scale and facilitating the discovery of digital phenotypes. As illustrated in Fig 1, the system is composed of a mobile health application and an open-source interactive data processing pipeline. To initiate the ChAMP System, the research administrator opens the ChAMP app and clicks start for each mood induction task when prompted. The ChAMP app is designed with simple screens indicating expected administrator actions (e.g. start, restart, save) and, large-font countdowns while within active tasks. The app collects continuous motion (via the onboard inertial measurement unit which captures acceleration and angular velocity at a sampling rate of 300 Hz) and audio data (via the onboard microphone at a sampling rate of 8 kHz), makes sounds to signal procedural requirements to the administrator and/or child, ends tasks, and stores organized data for researchers.

**TABLE 1.** Champ Behavior Battery Tasks

| Task | Approach | Speech | Bubbles |
| --- | --- | --- | --- |
| RDoC Construct | Potential Threat | Acute Threat | Reward |
| Duration | 30 s | 3 m | 3 m |
| Description | Child walks into darkened room toward covered object | Child tells story and is told it will be judged | Child plays with bubbles from a machine |
| Stimuli | Rapid beeping from 19 to 25 s to induce anxiety | Buzzer sounds at 90 and 160 s to induce startle | None |
| Output | Motion data as CSV | Audio data as mp3 | Motion data as CSV |

ChAMP App Behavioral Battery is composed of Approach, Speech and Bubbles mood-induction tasks which can be completed in under seven minutes. Each task addresses a research domain criteria (RDoC) construct associated with childhood mental health.

**Fig. 1.** To facilitate scalable validation and reproduction of the Kid Study we make the critical components to using the ChAMP mobile application widely accessible, including: a process for researchers to directly request ChAMP app download to enable their own data collection (left), the protocol for administering the ChAMP behavioral battery (middle), and an interactive open-source analysis script for reproducing biomarker feature sets (right).

The ChAMP App, written in Kotlin, was designed to be downloaded onto an Android smartphone and secured to the child’s lower back using a custom waist belt (belt instructions on the ChAMP Website [27]). The interactive data processing pipeline is implemented in Python in a Jupyter Notebook and automatically extracts features that quantify how a child moves and speaks during each task. Features are extracted from theory-driven phases within the task and provide a summary of movement and vocal data during each phase. Digital biomarkers are defined as features that demonstrate clinical relevance and can be combined to form digital phenotypes of childhood emotional and behavioral health. The platform enables seamless collaboration, so researchers with relevant skills can contribute new algorithms for the community to use.

### B. The ChAMP Assessment Battery

Each of the three mood induction tasks (see Table 1: Approach [18], [19], [28], Speech [20], [29], [30], and Bubbles [2], [3], [21]) induces an emotionally salient stimuli representing multiple constructs of daily functioning [31], associated with emotional and behavioral problems [6], [10], [32], [33]. Each task is described in detail in the full protocol (see [27]).

### C. Theory-Driven Feature Engineering

The Approach task was cut into six 5-second segments that capture each protocol sequence (i.e., walking into room; standing in front of covered object) and separate theorized threat response phases (i.e., potential threat, startle, and response modulation) [18], [34]. The Bubbles task was cut into twelve 15-second segments to explore potential changes in response to a single, repeated reward over time. As time until reward satiation is largely unexplored (outside of food reward) it was unknown whether or when reward satiation would be reached and so a span of seconds (similar, but somewhat longer than methodologies used in MRI reward studies [35]) was chosen, yet was largely exploratory. For these two tasks, angular velocity magnitude and acceleration magnitude were derived from the smartphone’s 3-axis gyroscope and accelerometer. For every segment, the median, sum, standard deviation, variance, and 5^th^, 25^th^, 75^th^, and 95^th^ percentiles of angular velocity and acceleration magnitudes were extracted to summarize the child’s movement within each segment. Additional details are available in [27].

The Speech task was cut into three segments: prior to the first buzzer (0-90 sec), between the first and second buzzer (90-150 sec), and after the second buzzer (150-180 sec). These segments attempt to capture differential aspects of fear including proactive avoidance of upcoming threat (being judged pre buzzers), and reactive escape from startle [36], as well as any changes due to habituation of a repeated stimuli [37] (after the second buzzer). Five seconds of audio at the beginning and end of the task and before and after each buzzer were removed to reduce the likelihood of any administrator audio being captured (i.e., giving instruction and/or time left). The WebRTC voice activity detector (VAD) was used to isolate the child’s voice in the recording for analysis [38] following which recording volume was increased by 15 dB to ensure we were accommodating for the posterior positioning of the phone. Audio features were selected from our previous work (e.g., (zero crossing rate (ZCR), Mel frequency cepstral coefficients (MFCC), spectral centroid [20]) and expanded to include proportion of time spent speaking in each task segment and throughout the whole task, chromagram (chroma stft), root mean square, spectral bandwidth, and the approximate minimum and maximum frequency. We also assessed the variance and mean pitch intensity, as well as the ranked list of dominant pitch intensities, all extracted using chroma stft [39]. We developed the ranked-dominant pitch intensity feature by summing the intensity of each pitch across the segment, and then ranking the pitches from 0-1 by the pitch intensity contribution, 0 being more dominant (e.g. mean ranked-dominant pitch intensity of Clll in diagnosed children was .727 versus for undiagnosed children it was .454, indicating the dominant pitch for undiagnosed children was more likely ranked higher for Clll than diagnosed children). Descriptive statistics (variance and/or mean) were computed for each feature. Additional details are available in [27].

### D. Study Recruitment and Experimental Protocol

Children and their primary caregiver were recruited as participants in Fall 2021-2022 from a state-wide sample via advertisements (n = 104) as well as in collaboration with the Vermont Center for Children, Youth and Families (VCCYF) in the Division of Child Psychiatry at the University of Vermont (n=6). To meet inclusion criteria, children had to be aged 4-8 years, with no suspected or diagnosed developmental disorders, serious medical conditions, or cognitive impairments, and primary caregivers had to be at least 18 years old and English-speaking.

For this analysis, 104 children between ages 4 and 8 years were included (M=81.42 months, SD=14.50), 61.5% (n=62) were male, and 89.4% (n=93) were White, non-Hispanic. Most reporting caregivers (80.2% biological mothers, n=77) were married (82.6%, n=71, 18 missing), had attained a bachelor’s degree or higher (88% n=75, 19 missing) and 81% (n=69, 19 missing) had average yearly household income of $75,000 or more. During the study lab visit, children were assessed for mental health diagnoses of which 47.6% (n=49) met criteria for at least one disorder of anxiety, depression or ADHD. Overall, 33% (n=34) had an anxiety disorder (inclusive of generalized anxiety disorder, separation anxiety disorder, social phobia, anxiety-unspecified, and post-traumatic stress disorder); 13.6%, (n=14) had a depressive disorder (inclusive of major depressive disorder, persistent depressive disorder and depressive disorder-unspecified); 19.4% (n=20) had a diagnosis of ADHD (inattentive, hyperactive or combined), including 11 children (10%) with an anxiety or depressive disorder (Fig 3). The range of disorders in this sample fit recommendations of recruiting participants with less specific clinical criteria, and without a “super-normal” control group to better represent general populations [40].

Eligible children and their primary caregivers attended a 2.5-hour laboratory visit. During the visit, the child’s caregiver was consented, administered a clinical interview about their child’s mental health (Schedule for Affective Disorders and Schizophrenia Preschool Version, present and lifetime) [41], [42] and completed several questionnaires about their child’s mental health and development. Diagnoses were derived via clinical consensus with a supervising licensed clinical psychologist using the best-estimate procedures based on parent-report symptom checklists and family history.

Simultaneously, the child was outfitted with sensors and a mobile phone (Fig 2A). The child was led to a novel room by an administrator to engage in the ChAMP Behavior Battery (Fig 2B). Afterward the child completed a structured intelligence test and played with toys until their caregiver completed their tasks. Families were compensated with a $60 gift card and a $5 toy. Families were called by a licensed clinical psychologist to discuss the results of their child’s mental health assessment and referral recommendations within two weeks of their visit. Study procedures were approved by the University of Vermont Institutional Review Board (CHRBSS 00001218).

**Fig. 2.**
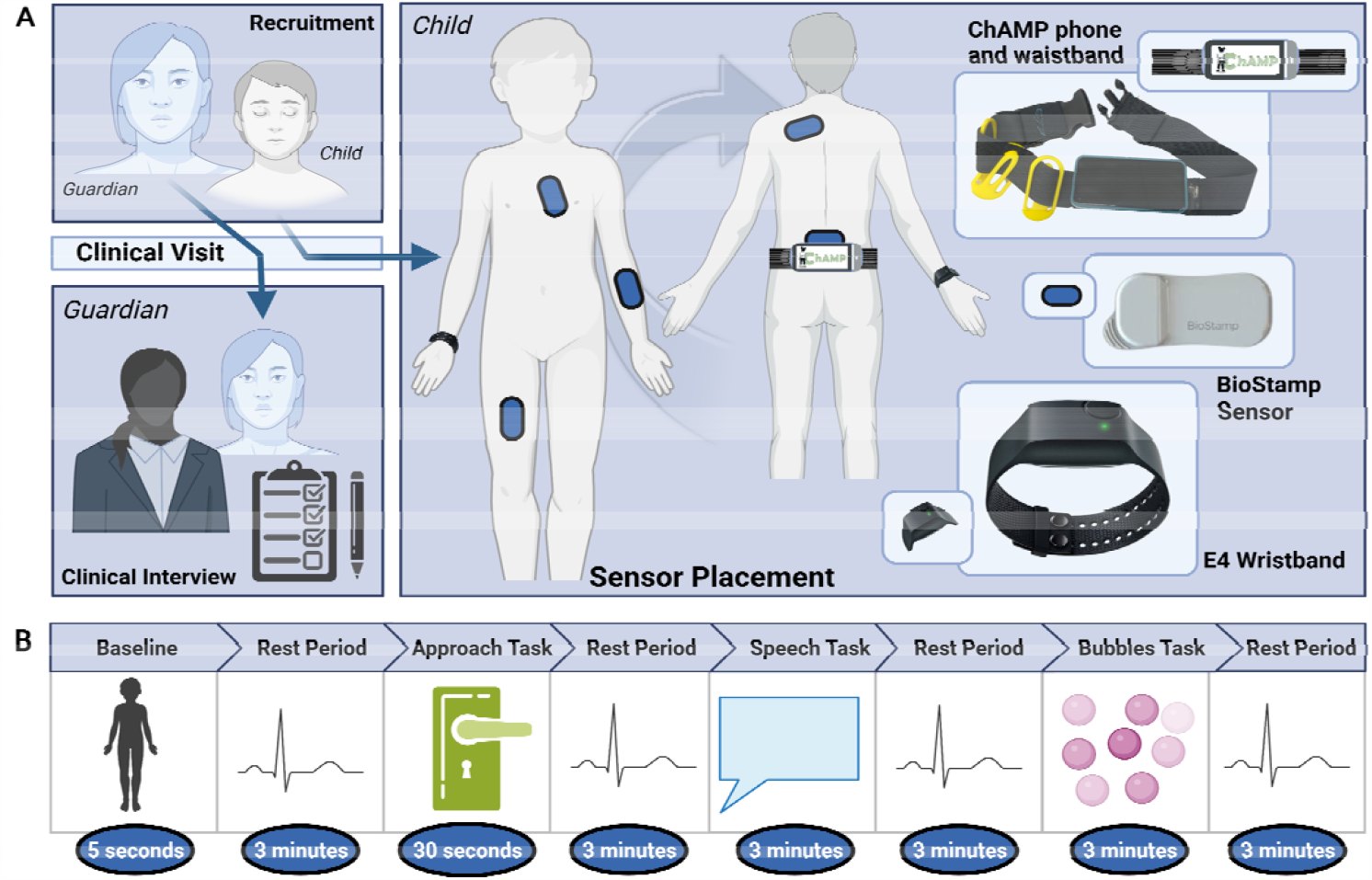
Participating children and their primary caregiver participate in a ∼2.5-hour study visit. The caregiver completes a clinical interview to assess their child’s mental health status (A, bottom left) while the child is outfitted with several wearable sensors including 5 BioStamp sensors, an E4 Empatica wrist sensor, and the belt-worn Android smartphone with the downloaded app, ChAMP (A, right) and completes the ChAMP behavioral assessment (B).

**Fig. 3.** Breakdown of participant diagnostic results by sex.

### E. Analytic Methods

Raw signal data needs context to interpret, and the theory driven phases used to segment each task provides this needed context. However, prior to assessment of theory driven phases, a determination of inclusion or exclusion of demographic information is needed for model feature selection. We performed t-tests to determine associations between age and disorder presence. We also used t-testing to assess if child sex was related to symptom severity for children who endorsed at least one symptom. We performed Chi squares testing to determine relationships between child sex and disorder presence. We then evaluated for correlations between age and symptom severity using Spearman correlations.

Preparing to dig deeper into understanding the relationship between the movement and audio features extracted from these tasks and emotional and behavioral health, we partitioned the dataset by task and by availability of data. This process produced 5 separate feature sets from the original 104 subjects, each were evaluated separately through model building efforts; Approach Task only (94 subjects), Bubbles Task only (99 subjects), Speech Task only (99 subjects), all three completed tasks (94 subjects), and 5-nearest neighbors imputation for tasks for subjects who were missing one or more tasks (101 subjects). Three subjects were not included in analysis as they had no sensor data. While in previous work we considered task-specific models, this is the first dataset we have considered that combined multiple valance tasks into one cohesive model.

To assess the potential for the ChAMP System to detect underlying mental health conditions in young children, we trained several binary classification models. Models were trained to identify children with depression, anxiety, or ADHD based on age, gender, and the 5 differing feature sets from the ChAMP System. To begin, we examine relative performance of Gaussian Naïve Bayes (NB), Support Vector Machine with Stochastic Gradient Descent Booster (SVM), Decision Tree (DT), XGBoost (XGB), and Logistic Regression (LR) classification models with varying infrastructures and feature selection methods. Specifically, we considered each classifier, with and without hyperparameter tuning, with and without decision thresholding weighted on balanced accuracy, trained on either 10 or 20 of the top predictors from 4 different feature importance methods. The feature importance methods considered ranked and selected predictors considering entropy-based Shannon information gain (DT), GINI impurity (GINI), permutation importance (PERM), and XGBoost feature importance (XGB). Models were trained and evaluated using stratified 2-fold shuffled cross-validation and characterized by the mean balanced accuracy (B-Accuracy), mean area under curve for the receiver operating curve (ROC AUC), sensitivity, specificity, false positive rate (FPR), and true positive rate (TPR) averaged across the two folds’ test cases. The primary metric defining performance ranking was balanced accuracy. Two folds were selected to maintain sufficient training instances for diagnoses that have small sample sizes (Fig 4C). For models predicting ADHD, there were 9-10 diagnosed individuals per fold. For anxiety, 15-16 diagnosed children per fold. For depression, there were 6 diagnosed children per fold. Evaluations of feature importance occurred within each fold of the cross validation. The feature set sizes of 10 and 20 were selected to limit model overfitting caused by too many considered features and to narrow down the modalities yielding the most influential biomarkers.

**Fig. 4.**
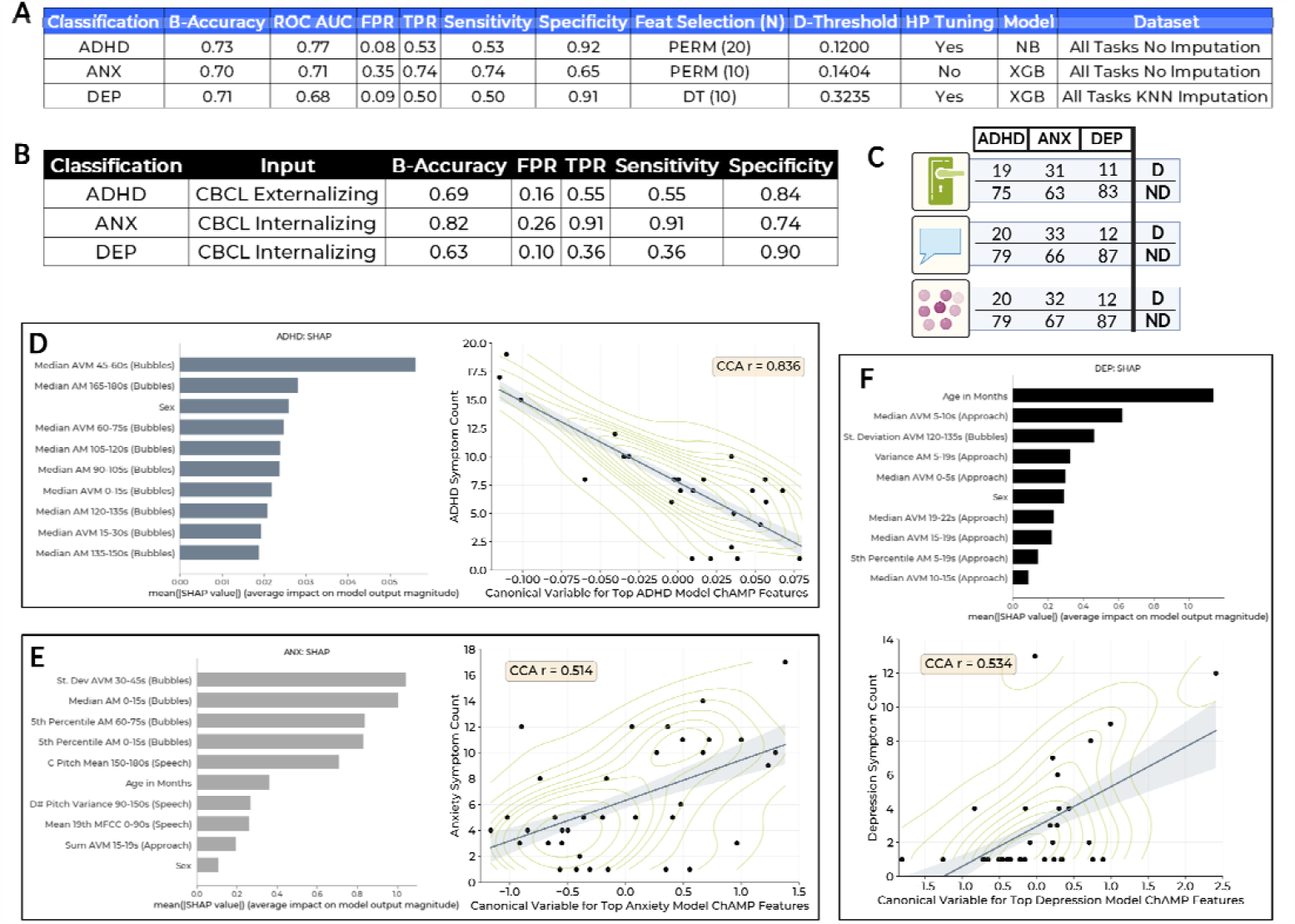
A. Highest performing binary diagnostic classification models for true clinical consensus prediction using ChAMP features and demographic features (sex and age in months). B. Parent report questionnaire (CBCL) concordance with true clinical consensus C. Breakdown of subject counts by task and diagnostic classification (A) for the Approach task (top, green door), Speech task (blue middle speech bubbles) and the Bubbles task (bottom pink bubble icon). Top 10 predictors as defined by Shapley Additive Explanations for overall model contributions in each binary diagnostic model for one fold of the highest performing model for ADHD (D right), Anxiety (E right), and Depression (F right). Canonical correlation plots for 1-component CCA comparing the canonical variable for the diagnostic predictors in each o the top moels compared to diagnostic symptom count for ADHD (D left), Anxiety (E left), and Depression (F left).NB = Naïve Bayes, XGB = XGBoost, PERM = Permutation Feature Importance, DT = Entropy-based Shannon information gain. FPR = False Positive Rate. TPR = True Positive Rate. D = Diagnosed. ND = Not Diagnosed. N = Number of Features Selected. D-Threshold = Decision Threshold Value. HP Tuning = Hyperparameter Tuning. ANX = Anxiety, DEP = Depression, B-Accuracy = Balanced Accuracy. KNN = K-Nearest Neighbors.

For models that were tuned, we selected a subset of parameters for each classifier to consider. For SVM, parameters of *learning_rate, loss, penalty, fit_intercept*, and *alpha* were tuned. For the DT classifier, *max_features, max_depth, criterion, min_samples_leaf*, and *min_samples_split*, were considered. For the NB classifier, we selected only *var_smoothing*. For XGB, *max_depth, min_child_weight, gamma, subsample*, and *reg_alpha*. For *LR, penalty, C*, and *class_weight* were tuned. Top performing models following model evaluations were then explored using SHAP (SHapley Additive exPlanations)[43] and compared to foundational work. Features identified in top performing models were then evaluated using one-component Canonical Correlation Analysis to assess the relationship between these top predictors and diagnostic symptom counts, a measure of symptom severity, for children who had at least one symptom.

## III. Results

### A. Demographic Associations with Diagnosis and Severity

Child age was not related to the presence of Anxiety (t(101)=.-1.88, p=.064) or ADHD (t(101)=.-.738, p=.462), but related to Depression (t(101)=.-2.88, p=.005) such that children with a diagnosis were older than those without. Child sex was unrelated to the presence of Anxiety (x(1,103)=.117, p=.732) ADHD (x(1,103)=.154, p=.695), and Depression (x(1,103)=.719, p=.397). For children with at least one symptom within diagnosis, child age was unrelated to symptom severity for Anxiety (r(41)=.089, p=.578), or ADHD (r(29)=-.018, p=.927), but was related to depression (r(38)=.356, p=.028). Child sex was related to symptom severity for Anxiety (t(39)=2.178, p=.036), such that females exhibited more symptoms than males, but child sex was unrelated to ADHD (t(27)=-.160, p=.874), or depression (t(37)=-.912, p=.368) symptoms. Based on several significant relationships found here, we included child sex and age in months in all model datasets.

### B. Predictive Modeling of Mental Health Diagnosis

We build several binary models classifying each diagnosis (anxiety, depression, ADHD) using only objective child physiology and behavior which would not require caregiver reporting that can be dependent on caregiver-child relationship, contexts for observations, caregiver mental health, mental health literacy, and societal stigma. All of the top performing models for each diagnostic classification (Fig 4A) favored feature sets that included features across all three behavioral tasks. Notably, each of these top models favored infrastructures that included decision thresholding for optimizing balanced accuracy and two of the three included hyperparameter tuning. All top performing models were able to correctly classify at least 50% of the true positives within the dataset and all models achieved balanced accuracy rates between 70 and 73%.

The top performing thresholds were all found to be lowered from the default rate (.5) to a threshold ranging from .12 to .32. This finding is interesting, as typically lowering a threshold so dramatically would cause an increased risk of false positives. However, the models we present still exhibit false positive rates in similar ranges to parent report (.08-.35 vs .1 to .26).

In previous work, we built several models considering the ability of features derived from each of the individual tasks in the ChAMP mood induction battery to classify the presence of an internalizing diagnosis (depression and/or anxiety) [21]. Sensitivities for top performing models for each task in previous work are similarly or less than the sensitivities achieved for classifying specific disorders (ADHD, depression, and anxiety) herein (.39-.67 vs .5-.77). For example, the model we present for anxiety has better sensitivity than any of the internalizing disorder models we have presented in prior work. Specificities of single-task models for internalizing disorders ranged from .64 to .93 in our prior work. In this work on multi-task diagnostic-specific models, have specificities ranging from .65 to .92.

### C. Evaluating Potential as a Complementary System

Consistent with our previous work, we compared psychometric evaluations of the questionnaire-based parent-reported CBCL with our smartphone-based movement and vocal feature models on child internalizing and externalizing diagnosis as determined via K-SADS-PL with clinical consensus. Clinical thresholds on the parent-report CBCL Externalizing Score detect ADHD with lower balanced accuracy (.69 vs. .73) and specificity (.84 vs. .92), but slightly higher sensitivity (.55 vs. .53) than models based on movement and vocal biomarkers (Fig 4A & B). Clinical thresholds on the parent-reported CBCL Internalizing Score for detecting depression exhibit lower balanced accuracy (.63 vs. .71), sensitivity (.36 versus .5), and specificity (.9 vs .91), higher false positive rate (.1 vs .09) and lower true positive rate (.36 versus .5) than models based on movement and vocal biomarkers (Fig 4A & B). Clinical thresholds on parent-reported CBCL Internalizing Score for detecting anxiety exhibited superior performance than the top performing ChAMP model in all metrics.

Our current and past studies have paralleled previous work demonstrating varied sensitivity of the CBCL, ranging from .0-.38 [44] to as high as .44 to .86 [45]. In our past work, biomarker-based models yielded a minimum 60% improvement in sensitivity over CBCL-based concordance to consensus report. In this work, we observed higher performance of CBCL-based concordance compared to previous work (.36-.91 vs. 00-.42 [23]) and thus sensitivities observed in ChAMP-based models did not yield such significant improvements relative to CBCL (.5-.74 vs .36-.91). The CBCL psychometrics observed here are similar to, and for some diagnostic groups higher than, those in larger studies [46,47]. Interestingly, anxiety in this sample of parents was reported with sensitivities higher than previously reported in any of our teams’ past work (.91). Notably, specificity of CBCL-based concordance in this study was similar to prior work (.88-1 [23] vs. .74-.90). ChAMP-based models we engineered performed with nearly parallel specificity to the observed CBCL-concordance in this study (.65-.92 vs. .74-.90).

In exploratory post-hoc analyses, we compared model false negatives (FNs) to true positives (TPs) on demographics (sex, age, race, income) and symptom characteristics (ADHD, depression and anxiety symptom counts and diagnostic subtypes), to investigate reasons the models may be missing some children with diagnoses. For ADHD, TPs were significantly younger than FPs (t(17)=-2.13, p=.049). For Depression, TPs may exhibit more ADHD symptoms than FPs (t(5.03)=2.10, p=.089), but did not reach statistical significance. For anxiety, there appeared to be no differences between TPs and FPs. Perhaps anxiety should be analyzed as subtypes (i.e., specific anxiety diagnoses, or fear vs. distress) in future analyses to improve model specificity. There were no significant differences between TNs and FNs for any diagnosis.

For CBCL internalizing reports, TPs exhibited more anxiety symptoms according to the KSADS interview than FP (t(38)=2.09, p=.043) and there were differences by group in terms of caregiver education. Caregivers with higher education were more likely to fall in the FP than the TP group (t(31)=-2.14, p=.040). For CBCL externalizing reports on KSADS ADHD diagnoses (which is not a direct comparison of ADHD), FPs were more likely to be boys than TPs (x(1,20)=3.78, p=.052) and there was a significant difference by group in terms of household income. Caregivers with higher income were more likely to be in the FP than the TP group (t(13)=-2.21, p=.045). Overall, analyses of subtypes may suggest that caregiver report may be influenced by caregiver demographics, and that physiological/behavioral models still need more analyses to understand better what is driving FPs.

We survey model explainability using Shapley Additive Explanations (SHAP)-based feature contributions [43]. All models selected features originating from more than one of the three behavioral tasks, rather than favoring a task-specific model. Interestingly, the ADHD model was the only of the top models to favor 20 instead of 10 features, and the top 10 features (Fig 4D) nearly all originated from the Bubbles task. The top performing Depression model included both of the two demographic features (Age and Sex), as well as features from both the Approach and Bubbles tasks. The Anxiety top performing model included both demographic features, as well as features across all three tasks.

### D. Child Diagnosis and Symptom Severity

Considering these top 10 to 20 features (depending on model), we performed Canonical Correlation Analysis (CCA) to evaluate the relationship between these predictors and symptom counts from the K-SADS-PL. We found that all predictor combinations yielded moderate to strong correlations (r = .51-.83) to diagnostic severity (defined by symptom count). This finding supports the hypothesis that theory-driven biomarkers may complement current screening by not only assisting in diagnostic classification, but also by potentially screening for estimated severity of an underlying disorder.

While these results are promising and show the potential of the ChAMP tool to collect valuable biomarkers for classifying childhood mental health status, significant future work is needed to engineer additional theory-driven features, integrate other physiological signals, and continue to explore modeling approaches to optimize performance. These results imply that each task in the ChAMP battery provides important, independent, and potentially clinically relevant, information for characterizing emotional and behavioral health that could complement parent report. Future work should also be done to evaluate how combinations of parent-report with wearable-sensor and smartphone-based signals may improve clinical capability by providing a complementary data stream for identifying and intervening on early childhood mental health concerns in young children.

## IV. Discussion

Through the introduction of the ChAMP system, we provide an accessible, digital tool to collect physiological and behavioral data during brief, structured mood induction tasks and an open-source platform to engineer objective features related to childhood emotional and behavioral health. We provide promising results suggesting that even some of the simplest movement and vocal features from these tasks could serve as biomarkers for childhood mental health.

### A. Relationship Within and Across Tasks

Our feature analyses demonstrate several key findings for future use. First, it is important to use a variety of multimodal features to model the complexity of child behavior during this battery. This implies that machine learning techniques which allow for modeling these complex relationships will likely be the best analytical strategy for these data [1], [19], [20]. Herein, we used only high-level features. However, based on our previous work, it is likely that that more granular biomechanical features may provide improved model performance [23]. Thus, we aim to explore a greater variety and complexity of features (e.g., turning behavior, postural sway [48]) in future work. Second, engineering features from temporal segments within each task is crucial as results revealed that specific task segments are differentially important in classifying children with and without a disorder (Fig 4D-F). While behaviorally coded features from mood induction tasks are typically summed across one (or multiple) tasks [3], [10], our task protocols and measurements were developed and adapted to evoke and quantify differential magnitudes of emotional stimuli across time. Thus, including all three mood induction tasks as one behavior battery is important as features from each appear to contribute uniquely to disorder classification. These findings are important in the context of our past work, as we have never before combined results across mood-induction tasks.

### B. Association with Diagnoses and Symptom Counts

In our prior work in a separate sample, children with a diagnosis turned away more and more quickly from an unknown threat in the Approach task compared to children without a diagnosis [18]. We observed this phase showed importance in this new sample as well, especially for the classification of Depression and Anxiety. These heightened features (herein, median and sum of movement magnitude, and previously, more tilt/turn of the child’s body) could represent biomarkers of elevated attentiveness and responsiveness to potential threat, consistent with work demonstrating children with internalizing disorders attended more to potentially threatening faces than those without [33]. Second, we found higher dominant pitch tone after an unexpected startle in the Speech task in children with an Anxiety diagnosis, consistent with our previous work in a separate sample [20]. These heightened biomarkers represent elevated reactivity to acute threat, consistent with work demonstrating adults with fear-related anxiety disorders startle more (higher magnitude eye blink) than those without [49]. Third, we found that the Bubbles task was dominantly representative across top features in the diagnostic prediction models. This supports our previous work demonstrating that the Bubbles task biomarkers were more related to broad emotional dysregulation than specific diagnostic symptoms [21].

We have extended our past work by implementing a variety of supervised classification models which further demonstrate the importance of including multi-modal features across temporal phases, and tasks as multiple modalities, phases, and tasks were included in each optimal diagnostic model. These models perform on-par with, or improve upon, several of our previous models presented in foundational efforts and incorporate several mood induction tasks in a structured battery. We also demonstrate, for the first time, that linear combinations of top-performing ChAMP features during these tasks are strongly associated with symptom counts of child anxiety, depression, and, most notably, ADHD. Future work should consider additional wearable sensors, features, subjects, and modeling approaches (e.g., deep learning or transfer learning).

A strength of this study is the inclusion of a wide range of internalizing disorders, while not excluding externalizing disorders. This allowed us to advance modelling efforts to specialize classification to specific diagnoses under the internalizing and externalizing umbrella, which we previously had not evaluated. This may make our small sample more clinically representative of community populations. Overall, findings show promise that specific segments of each of the three mood induction tasks may contribute to future models for detecting childhood anxiety and depression and for assessing severity of underlying diagnosis.

It is also important to note potential limitations relating to bias. We have found that our models less clearly portray biases related to demographic background in parent-report surveys on child mental health status, however we cannot affirm that the models built are without bias. Notably, our sample may not represent the general population in terms of demographics or impairments. Our sample was less racially and ethnically diverse, and more affluent than that US population. Families of children with impairment appeared more likely than those without to self-select into the study. During recruitment, community waitlists for diagnostic evaluations were long (>9 months) and receiving a diagnostic assessment for their child within two weeks through participation in the study was reported as a significant motivator for families. It is also possible that the emotional impact of COVID anxiety/isolation may have increased child impairment as seen in adolescent samples[50]. We aim in future work to further bolster our training samples with a wider diversity of children and families.

### C. ChAMP as an Open-Access Digital Tool

In the past decade, we have seen a rise in validated mobile applications and platforms that aim to increase awareness, monitoring, and intervention of mental illness (e.g., PanicMechanic [51],[52], LAMP [25], FOCUS [53]). There is an increase in commercial and research platforms that incorporate aspects of the digital phenotyping lifecycle (*i*.*e*., study protocol portal, data collection tool, mechanism for storing and accessing data, data analysis tools), but very few feature all of the necessary components and data to replicate and validate findings [15]. ChAMP provides accessibility and flexibility in these components, and for the first time, with a focus on early childhood. We hope with this platform that other researchers may extend this work to explore intrasubject variability and repeatability of findings across additional participant groups.

By offering an app and platform, we aim to aid researchers in advancing the field of digital psychiatry [54]. We aim to increase explainability for this toolkit, and accessibility for approved clinical researchers utilizing the toolkit, by providing a website with additional documentation [27]. Future improvements to ChAMP will increase the customizability of our platform to enhance research and future clinical utility [15], [54]. Our next steps toward a customizable tool would allow investigation into additional conditions impacting children across a wider range of ages. Potential modifications could include additional mood induction tasks, project-specific enhancements to app protocols, and companion wearable devices for enabling additional sensing modalities. For instance, autism is an area where movement and speech data has often been used for assessment of diagnostic status and severity [55], [56] as well as ADHD [57], [58]. With more replication and validation across samples by clinical researchers, precision and model performance can be further enhanced. This is important, as a future iteration of a similar objective system must have high accuracy so as to not overburden the system or family with false positive reports or under-identify children that may be struggling un-noticed. Moreover, this system, once further refined, could have potential to ultimately improve assessment by serving as a complementary clinical endpoint to better inform intervention.

## Data Availability

A subset of the movement data derived from the Approach and Bubbles tasks used in this analysis can be downloaded from our website: ChAMP Website). The diagnostic data associated with each subject is included in a summarized format.

https://brynchristineloftness.github.io/CHAMP/intro.html

## Acknowledgment

This work would not be possible without the wonderful participating families and support of the undergraduate research assistants conducting lab visits. All figures were created using Biorender.com.

## References

[1] E. W. McGinnis, W. Copeland, L. Shanahan, and H. L. Egger, “Parental perception of mental health needs in young children,” Child Adolesc. Ment. Health, vol. 27, no. 4, pp. 328–334, Nov. 2022, doi: 10.1111/camh.12515.

[2] S. Hurwitz and N. Yirmiya, “Autism diagnostic observation schedule (ADOS) and its uses in research and practice,” Compr. Guide Autism, pp. 345–353, 2014.

[3] J. R. Gagne, C. A. Van Hulle, N. Aksan, M. J. Essex, and H. H. Goldsmith, “Deriving childhood temperament measures from emotion-eliciting behavioral episodes: scale construction and initial validation,” Psychol. Assess., vol. 23, no. 2, pp. 337–353, Jun. 2011, doi: 10.1037/a0021746.

[4] C. Lord et al., “The autism diagnostic observation schedule-generic: a standard measure of social and communication deficits associated with the spectrum of autism,” J. Autism Dev. Disord., vol. 30, no. 3, pp. 205–223, Jun. 2000.

[5] A. C. Sousa, S. N. Ferrinho, and B. Travassos, “The Use of Wearable Technologies in the Assessment of Physical Activity in Preschool- and School-Age Youth: Systematic Review,” Int. J. Environ. Res. Public. Health, vol. 20, no. 4, Art. no. 4, Jan. 2023, doi: 10.3390/ijerph20043402.

[6] L. M. McTeague and P. J. Lang, “The anxiety spectrum and the reflex physiology of defense: from circumscribed fear to broad distress,” Depress. Anxiety, vol. 29, no. 4, pp. 264–281, Apr. 2012, doi: 10.1002/da.21891.

[7] C. E. Durbin, E. P. Hayden, D. N. Klein, and T. M. Olino, “Stability of laboratory-assessed temperamental emotionality traits from ages 3 to 7,” Emot. Wash. DC, vol. 7, no. 2, pp. 388–399, May 2007, doi: 10.1037/1528-3542.7.2.388.

[8] A. M. Kring and D. M. Sloan, “The Facial Expression Coding System (FACES): development, validation, and utility.,” Psychol. Assess., vol. 19, no. 2, p. 210, 2007.

[9] H. H. Goldsmith and M. K. Rothbart, “The laboratory temperament assessment battery (LAB-TAB),” Univ. Wis., 1993.

[10] J. S. Moser, C. E. Durbin, C. J. Patrick, and N. B. Schmidt, “Combining Neural and Behavioral Indicators in the Assessment of Internalizing Psychopathology in Children and Adolescents,” J. Clin. Child Adolesc. Psychol. Off. J. Soc. Clin. Child Adolesc. Psychol. Am. Psychol. Assoc. Div. 53, Jan. 2014, doi: 10.1080/15374416.2013.865191.

[11] G. Kochanska, K. Murray, and K. C. Coy, “Inhibitory control as a contributor to conscience in childhood: from toddler to early school age,” Child Dev., vol. 68, no. 2, pp. 263–277, Apr. 1997.

[12] K. A. Callender, S. L. Olson, D. C. R. Kerr, and A. J. Sameroff, “Assessment of Cheating Behavior in Young School-Age Children: Distinguishing Normative Behaviors From Risk Markers of Externalizing Psychopathology,” J. Clin. Child Adolesc. Psychol., vol. 39, no. 6, pp. 776–788, Nov. 2010, doi: 10.1080/15374416.2010.517165.

[13] L. S. Wakschlag et al., “Defining the ‘disruptive’ in preschool behavior: what diagnostic observation can teach us,” Clin. Child Fam. Psychol. Rev., vol. 8, no. 3, pp. 183–201, Sep. 2005, doi: 10.1007/s10567-005-6664-5.

[14] G. Yáñez-Téllez et al., “Cognitive and executive functions in ADHD,” Actas Esp. Psiquiatr., vol. 40, no. 6, pp. 293–298, Nov. 2012.

[15] J. Torous, M. V. Kiang, J. Lorme, and J.-P. Onnela, “New Tools for New Research in Psychiatry: A Scalable and Customizable Platform to Empower Data Driven Smartphone Research,” JMIR Ment. Health, vol. 3, no. 2, p. e16, May 2016, doi: 10.2196/mental.5165.

[16] S. Perochon et al., “A scalable computational approach to assessing response to name in toddlers with autism,” J. Child Psychol. Psychiatry, vol. 62, no. 9, pp. 1120–1131, 2021, doi: 10.1111/jcpp.13381.

[17] B. C. Loftness, J. Halvorson-Phelan, A. O’Leary, N. Cheney, E. W. McGinnis, and R. S. McGinnis, “UVM KID Study: Identifying Multimodal Features and Optimizing Wearable Instrumentation to Detect Child Anxiety,” in 2022 44th Annual International Conference of the IEEE Engineering in Medicine & Biology Society (EMBC), Jul. 2022, pp. 1141–1144. doi: 10.1109/EMBC48229.2022.9871090.

[18] E. W. McGinnis et al., “Movements Indicate Threat Response Phases in Children at Risk for Anxiety,” IEEE J. Biomed. Health Inform., vol. 21, no. 5, pp. 1460–1465, Sep. 2017, doi: 10.1109/JBHI.2016.2603159.

[19] E. W. McGinnis et al., “Wearable sensors detect childhood internalizing disorders during mood induction task,” PloS One, vol. 13, no. 4, p. e0195598, 2018, doi: 10.1371/journal.pone.0195598.

[20] E. W. McGinnis et al., “Giving Voice to Vulnerable Children: Machine Learning Analysis of Speech Detects Anxiety and Depression in Early Childhood,” IEEE J. Biomed. Health Inform., vol. 23, no. 6, pp. 2294–2301, Nov. 2019, doi: 10.1109/JBHI.2019.2913590.

[21] E. W. McGinnis et al., “Digital Phenotype for Childhood Internalizing Disorders: Less Positive Play and Promise for a Brief Assessment Battery,” IEEE J. Biomed. Health Inform., vol. 25, no. 8, pp. 3176–3184, Aug. 2021, doi: 10.1109/JBHI.2021.3053846.

[22] B. C. Loftness et al., “Toward Digital Phenotypes of Early Childhood Mental Health via Unsupervised and Supervised Machine Learning,” Feb. 2023, 10.1101/2023.02.24.23286417.

[23] R. S. McGinnis et al., “Rapid detection of internalizing diagnosis in young children enabled by wearable sensors and machine learning,” PLOS ONE, vol. 14, no. 1, p. e0210267, Jan. 2019, doi: 10.1371/journal.pone.0210267.

[24] S. Lagan et al., “Mental Health App Evaluation: Updating the American Psychiatric Association’s Framework Through a Stakeholder-Engaged Workshop,” Psychiatr. Serv. Wash. DC, vol. 72, no. 9, pp. 1095–1098, Sep. 2021, doi: 10.1176/appi.ps.202000663.

[25] J. Torous et al., “Creating a Digital Health Smartphone App and Digital Phenotyping Platform for Mental Health and Diverse Healthcare Needs: an Interdisciplinary and Collaborative Approach,” J. Technol. Behav. Sci., vol. 4, no. 2, pp. 73–85, Jun. 2019, doi: 10.1007/s41347-019-00095-w.

[26] H. Wisniewski, P. Henson, and J. Torous, “Using a Smartphone App to Identify Clinically Relevant Behavior Trends via Symptom Report, Cognition Scores, and Exercise Levels: A Case Series,” Front. Psychiatry, vol. 10, p. 652, 2019, doi: 10.3389/fpsyt.2019.00652.

[27] “ChAMP Homepage — ChAMP, A Scalable Mental Health Screening App Using Mood Induction Tasks.” https://brynchristineloftness.github.io/CHAMP/intro.html (accessed Jun. 01, 2023).

[28] N. L. Lopez-Duran, N. J. Hajal, S. L. Olson, B. T. Felt, and D. M. Vazquez, “Individual differences in cortisol responses to fear and frustration during middle childhood,” J. Exp. Child Psychol., vol. 103, no. 3, pp. 285–295, Jul. 2009, doi: 10.1016/j.jecp.2009.03.008.

[29] P. C. Kendall, E. Flannery-Schroeder, S. M. Panichelli-Mindel, M. Southam-Gerow, A. Henin, and M. Warman, “Therapy for youths with anxiety disorders: a second randomized clinical trial,” J. Consult. Clin. Psychol., vol. 65, no. 3, pp. 366–380, Jun. 1997.

[30] Y. J. Bae et al., “The hyporeactivity of salivary cortisol at stress test (TSST-C) in children with internalizing or externalizing disorders is contrastively associated with α-amylase,” J. Psychiatr. Res., vol. 71, pp. 78–88, Dec. 2015, doi: 10.1016/j.jpsychires.2015.09.013.

[31] “Research Domain Criteria (RDoC),” National Institute of Mental Health (NIMH). https://www.nimh.nih.gov/research/research-funded-by-nimh/rdoc (accessed Dec. 14, 2022).

[32] I. Baji et al., “[Symptoms of depression in children and adolescents in relation to psychiatric comorbidities],” Psychiatr. Hung. Magy. Pszichiátriai Társ. Tudományos Folyóirata, vol. 27, no. 2, pp. 115–126, 2012.

[33] G. A. Salum et al., “Threat bias in attention orienting: evidence of specificity in a large community-based study,” Psychol. Med., vol. 1, no. 1, pp. 1–13, 2012.

[34] J. J. Gross, “Emotion regulation: affective, cognitive, and social consequences,” Psychophysiology, vol. 39, no. 3, pp. 281–291, May 2002, doi: 10.1017.S0048577201393198.

[35] J. P. O’Doherty, “Reward representations and reward-related learning in the human brain: insights from neuroimaging,” Current Opinion in Neurobiology, vol. 14, no. 6, pp. 769–776, Dec. 2004, 10.1016/j.conb.2004.10.016.

[36] J. E. LeDoux, J. Moscarello, R. Sears, and V. Campese, “The birth, death and resurrection of avoidance: a reconceptualization of a troubled paradigm,” Molecular Psychiatry, vol. 22, no. 1, pp. 24–36, Oct. 2016, 10.1038/mp.2016.166.

[37] C. T. Sege, D. L. Taylor, J. W. Lopez, H. Fleischmann, E. J. White, and L. M. McTeague, “Coping in the Clinic: Effects of Clinically Elevated Anxiety on Dynamic Neurophysiological Mechanisms of Escape/Avoidance Preparation,” Biological Psychiatry: Cognitive Neuroscience and Neuroimaging, Aug. 2022, 10.1016/j.bpsc.2022.07.010.

[38] J. Wiseman, “wiseman/py-webrtcvad.” Dec. 15, 2022. Accessed: Dec. 16, 2022. [Online]. Available: https://github.com/wiseman/py-webrtcvad

[39] “librosa.feature.chroma_stft — librosa 0.10.0.dev0 documentation.” https://librosa.org/doc/main/generated/librosa.feature.chroma_stft.htm l (accessed Dec. 16, 2022).

[40] B. N. Cuthbert and T. R. Insel, “Toward the future of psychiatric diagnosis: the seven pillars of RDoC,” BMC Med., vol. 11, p. 126, 2013, doi: 10.1186/1741-7015-11-126.

[41] B. Birmaher et al., “Schedule for Affective Disorders and Schizophrenia for School-Age Children (K-SADS-PL) for the Assessment of Preschool Children-A Preliminary Psychometric Study,” J. Psychiatr. Res., vol. 43, no. 7, pp. 680–686, Apr. 2009, doi: 10.1016/j.jpsychires.2008.10.003.

[42] J. Kaufman, B. Birmaher, D. A. Brent, N. D. Ryan, and U. Rao, “K-SADS-PL,” J. Am. Acad. Child Adolesc. Psychiatry, vol. 39, no. 10, p. 1208, Oct. 2000, doi: 10.1097/00004583-200010000-00002.

[43] S. M. Lundberg and S.-I. Lee, “A Unified Approach to Interpreting Model Predictions,” Neural Information Processing Systems, 2017. https://papers.nips.cc/paper_files/paper/2017/hash/8a20a8621978632d76c43dfd28b67767-Abstract.html

[44] C. W. Rishel, C. Greeno, S. C. Marcus, M. K. Shear, and C. Anderson, “Use of the Child Behavior Checklist as a Diagnostic Screening Tool in Community Mental Health,” Research on Social Work Practice, vol. 15, no. 3, pp. 195–203, May 2005, 10.1177/1049731504270382.

[45] Aschenbrand SG, Angelosante AG, Kendall PC. Discriminant validity and clinical utility of the CBCL with anxiety-disordered youth. J Clin Child Adolesc Psychol. 2005;34: 735–746. pmid:16232070

[46] de la Osa N, Granero R, Trepat E, Domenech JM, Ezpeleta L. The discriminative capacity of CBCL/1½-5-DSM5 scales to identify disruptive and internalizing disorders in preschool children. Eur Child Adolesc Psychiatry. 2016; 25: 17–23. 10.1007/s00787-015-0694-4

[47] DS, Mogg K, Bradley BP, Montgomery L, Monk CS, McClure E, et al. Attention Bias to Threat in Maltreated Children: Implications for Vulnerability to Stress-Related Psychopathology. AJP. 2005; 162:291–296. 10.1176/appi.ajp.162.2.291 PMID: 15677593

[48] B. M. Meyer et al., “How Much Data Is Enough? A Reliable Methodology to Examine Long-Term Wearable Data Acquisition in Gait and Postural Sway,” Sensors, vol. 22, no. 18, Art. no. 18, Jan. 2022, doi: 10.3390/s22186982.

[49] L. M. McTeague and P. J. Lang, “The anxiety spectrum and the reflex physiology of defense: from circumscribed fear to broad distress,” Depress. Anxiety, vol. 29, no. 4, pp. 264–281, Apr. 2012, doi: 10.1002/da.21891.

[50] S. Wang et al., “Depression and anxiety among children and adolescents pre and post COVID-19: A comparative meta-analysis,” Frontiers in Psychiatry, vol. 13, p. 917552, Aug. 2022, 10.3389/fpsyt.2022.917552.

[51] R. S. McGinnis, E. W. McGinnis, C. Petrillo, J. Ferri, J. Scism and M. Price, “Validation of Smartphone Based Heart Rate Tracking for Remote Treatment of Panic Attacks,” in IEEE Journal of Biomedical and Health Informatics, vol. 25, no. 3, pp. 656–662, March 2021, doi: 10.1109/JBHI.2020.3001573.

[52] E. McGinnis, A. O’Leary, R. Gurchiek, W. Copeland, and R. McGinnis, “PanicMechanicTM: A Digital Therapeutic Delivering Biofeedback for Panic Attacks (Preprint),” JMIR Formative Research, Aug. 2021, 10.2196/32982.

[53] Ben-Zeev D, Kaiser SM, Brenner CJ, Begale M, Duffecy J, Mohr DC. Development and Usability Testing of FOCUS: A Smartphone System for Self-Management of Schizophrenia. Psychiatr Rehabil J. 2013;36(4):289–296. doi:10.1037/prj0000019

[54] McGinnis RS, McGinnis EW. Advancing Digital Medicine with Wearables in the Wild. Sensors. 2022;22(12):4576. doi:10.3390/s22124576

[55] Mohanta A, Mukherjee P, Mirtal VK. Acoustic Features Characterization of Autism Speech for Automated Detection and Classification. In: 2020 National Conference on Communications (NCC). ; 2020:1–6. doi:10.1109/NCC48643.2020.9056025

[56] Siddiqui UA, Ullah F, Iqbal A, et al. Wearable-Sensors-Based Platform for Gesture Recognition of Autism Spectrum Disorder Children Using Machine Learning Algorithms. Sensors. 2021;21(10):3319. doi:10.3390/s21103319

[57] Jiang X, Chen Y, Huang W, et al. WeDA: Designing and Evaluating A Scale-driven Wearable Diagnostic Assessment System for Children with ADHD. In: Proceedings of the 2020 CHI Conference on Human Factors in Computing Systems. CHI ‘20. Association for Computing Machinery; 2020:1–12. doi:10.1145/3313831.3376374

[58] Luo J, Huang H, Wang S, et al. A Wearable Diagnostic Assessment System vs. SNAP-IV for the auxiliary diagnosis of ADHD: a diagnostic test. BMC Psychiatry. 2022;22(1):415. doi:10.1186/s12888-022-04038-3

